# Costs of hand hygiene for all in household settings - estimating the price tag for the 46 least developed countries

**DOI:** 10.1101/2021.08.16.21262011

**Authors:** Ian Ross, Joanna Esteves Mills, Tom Slaymaker, Richard Johnston, Guy Hutton, Robert Dreibelbis, Maggie Montgomery

## Abstract

**Introduction:** Domestic hand hygiene could prevent over 500,000 attributable deaths per year, but 6 in 10 people in least developed countries (LDCs) do not have a handwashing facility with soap and water available at home. We estimated the economic costs of universal access to basic hand hygiene services in household settings in 46 LDCs.

**Methods:** Our model combines quantities of households with no handwashing facility (HWF) and prices of promotion campaigns, HWFs, soap, and water. For quantities, we used estimates from the WHO/UNICEF Joint Monitoring Programme. For prices, we collated data from recent impact evaluations and electronic searches. Accounting for inflation and purchasing power, we calculated costs over 2021-2030, and estimated total cost probabilistically using Monte Carlo simulation.

**Results:** An estimated US$ 12.2 - 15.3 billion over 10 years is needed for universal hand hygiene in household settings in 46 LDCs. The average annual cost of hand hygiene promotion is $334 million (24% of annual total), with a further $233 million for ‘top-up’ promotion (17%). Together, these promotion costs represent $0.47 annually per head of LDC population. The annual cost of HWFs, a purpose-built drum with tap and stand, is $174 million (13%). The annual cost of soap is $497 million (36%), and water $127 million (9%).

**Conclusion:** The annual cost of behaviour change promotion to those with no handwashing facility represents 4.7% of median government health expenditure in LDCs, and 1% of their annual aid receipts. These costs could be covered by mobilising resources from across government and partners, and could be reduced by harnessing economies of scale and integrating hand hygiene with other behaviour change campaigns where appropriate. Innovation is required to make soap more affordable and available for the poorest households.

**Summary box:** *What is already known?:*

- Understanding resource requirements is important for planning, but data on the costs of improving domestic hand hygiene are scarce.
- While a 2016 study estimated the global cost of drinking water, sanitation and hygiene, it did not report hygiene-specific estimates of recurrent or total cost, nor did it describe the assumed promotion intervention and handwashing facility or consider alternatives to them.

*What are the new findings?:*

- The total economic cost over 10 years is US$ 12.2 – 15.3 billion, of which $4.9 – 6.6 billion (42%) is for behaviour change promotion interventions.
- The remainder is for facilities and supplies, with soap the biggest cost category (36%) followed by handwashing facilities (13%) and water (9%).
- The facility and supply costs per household comprise an initial investment in a handwashing facility (lasting 5 years) at a median of US$ 17, accompanied by an annual cost of $17 for soap and $5 for water.

*What do the new findings imply?:*

- The annual cost of behaviour change promotion to those with no handwashing facility represents 4.7% of median government health expenditure in LDCs.
- On top of this, investments in infrastructure and supplies are required. Soap in particular is a substantial and recurrent cost, which may be unaffordable for the poorest households.
- Promotion costs could be covered by mobilising resources from across government and partners, and could be reduced by harnessing economies of scale and integrating hand hygiene with other behaviour change campaigns where appropriate.

## 1. Introduction

Hand hygiene reduces transmission of a variety of enteric and respiratory infections.^1,2^ Every year, 165,000 deaths from diarrhoeal disease and 370,000 deaths from acute respiratory infections are attributable to inadequate hand hygiene.^3^ However, nearly a third of the global population do not have handwashing facilities with soap and water available at home, denoted a “basic” hygiene service.^4^ Many more do not practice handwashing with soap at critical times – for example, only 26% of potential faecal contact events globally are followed by handwashing with soap.^5^ The COVID-19 pandemic highlighted the need for hand hygiene to reduce transmission across settings, including households, schools, healthcare facilities, and public places.^6^ The greatest deficit is in least developed countries (LDCs), where 6 in 10 people are without a basic hygiene service, of which about half have a handwashing facility but no soap and/or water.^4^ Hand hygiene is best facilitated by an on-premises water supply,^7^ but 60% of the LDC population do not have such a service,^4^ instead hauling water from off-premises sources.

Reviews of factors for success in scaling up public health interventions in low- and middle-income countries (LMICs) frequently identify costing and economic analysis of interventions in the top three success factors.^8,9^ For hand hygiene, such cost figures are scarce. A 2016 study costing the sustainable development goal (SDG) targets for drinking water, sanitation and hygiene (WASH) in 140 countries by Hutton and Varughese identified only five peer-reviewed studies providing data on the costs of handwashing promotion, alongside seven from grey literature.^10,11^ The absence of robust cost data for handwashing programmes makes financial planning and resource allocation difficult.^12^

Hutton and Varughese reported an annual capital cost of US$ 2.0 billion over 15 years for 140 LMICs to achieve basic hand hygiene in domestic settings.^10,11^ This figure includes handwashing facilities (HWFs) and promotion only. Limitations of these cost estimates include: (i) not reporting hygiene-specific estimates of recurrent or total costs, only incorporating them into totals for WASH; (ii) no description of the assumed hygiene promotion “software” or the assumed HWF type(s); (iii) inconsistent approach to HWF useful life, with 10 years applied for rural areas in all countries, but 2.5 or 5 years for urban areas in 86% of countries; (iv) no sensitivity analysis specific to hygiene assumptions.

The costs of activities promoting hand hygiene in LDCs are typically borne by governments and donors as a public health investment. However, only 16% of the 115 countries responding to a WHO-led survey could report the size of hygiene budgets or expenditures, compared to 53% for sanitation and/or water supply.^13^ Of the 16 countries providing expenditure data, 8 indicated only one source of funding (government, households, or donors), indicating that the data are not comprehensive.^13^ While the costs of HWFs, soap, and water are borne by households in the majority of cases, they can also be subsidised directly or indirectly (e.g. specific subsidies, cash transfers or humanitarian response). Affordability is a concern, and survey data points to a strong socio-economic gradient in soap availability within households.^14^

In this study, we aim to estimate the economic costs of universal access to basic hand hygiene services in household settings in 46 LDCs. Underlying objectives were to facilitate discussions and plans at the national and global levels, especially in light of the ongoing COVID-19 pandemic, and evaluate the size of the cost in relation to other investment priorities. We build on the Hutton and Varughese study by undertaking a thorough search for more price data, describing the assumed intervention and HWF, considering alternative scenarios for these, and characterising the uncertainty deriving from hygiene-related assumptions and data.

## 2. Methods

We built a model combining quantities of targeted households with prices of capital and recurrent items, to estimate the economic costs of basic hand hygiene in domestic settings in 46 LDCs (2021 list – Supplementary Material A).^15^ We followed the reference case of the Global Health Cost Consortium.^12^ The model structure is visualised in Supplementary Material B. Ethical approval was not required because the study analysed secondary data from published studies.

### Costing perspective and approach

We estimated economic costs from a societal perspective, and address who might bear those costs in the discussion section. We model straight-line scale-up of a hand hygiene promotion intervention (described below) over a 10-year horizon (2021-2030), whereby 10 equal cohorts of unserved households per country receive the intervention per year. Each cohort starts incurring recurrent costs in the year they receive the intervention. The scope of costed inputs comprised all activities contributing to behaviour change and purchase/use of a HWF over its useful life. We analysed quantities and prices per country for the 46 LDCs, separately for urban/rural areas. We then aggregated to an LDC total. Following norms in resource requirement estimation, we estimate the cost of reaching all target households, and do not incorporate the effectiveness of interventions.^12^

### Data on quantities

We retrieved hygiene service level estimates for the 46 LDCs from the WHO/UNICEF Joint Monitoring Programme (JMP).^4^ For countries missing JMP estimates for 2020 (Supplementary Material B), we applied an earlier year wherever possible (6 countries). In the absence of any data we applied the LDC average (5 countries). We calculated the number of households to be targeted per country by urban and rural setting, based on: (i) JMP coverage estimates; (ii) average household sizes for urban/rural from the latest Demographic and Health Survey;^16^ (iii) UN medium-variant population projections;^17^ (iv) assuming one HWF per household (equation in Supplementary Material B).

For our headline result (A), the population for which interventions are costed comprises households with “no handwashing facility”.^4^ This is because the relevant SDG indicator focuses on universal access to a HWF with soap and water on premises, rather than hygiene behaviour. However, we separately estimate two other results: (B) soap and water costs for households with a “limited” service (HWF observed but soap and/or water missing); (C) promotion costs if the whole LDC population is targeted regardless of JMP status. The latter may be required because, even though handwashing with soap is twice as likely to be practised when a designated handwashing facility is present,^5^ overall prevalence remains low. For example, it is estimated that in high-income countries, where over 99% of the population have water accessible on premises,^4^ only 51% of faecal contacts are followed by handwashing with soap.^5^ It may also be more practical to deliver behaviour change promotion to whole communities, rather than targeting on the basis of HWF ownership. Our headline result would, then, underestimate promotion costs if populations beyond those without HWFs are to be targeted.

There were 61 million households in LDCs with “no hygiene service” in 2020 (71% of which are in rural areas), and a further 81 million with a “limited” service (69% in rural areas). We used JMP analysis of the latest household survey containing HWF observation data for 42 of the 46 LDCs (Rick Johnston, personal communication). We divided a country’s population with a “limited” service into three categories based on the limiting factor(s): missing soap only (47% on average), missing water only (10%) and missing both (43%). We attribute the appropriate recurrent costs for those in the “limited” group based on the split per country.

### Data on prices

We collated price data from studies included in an earlier review,^10^ electronic searches (Supplementary Material B), and by contacting impact evaluation investigators. For the latter, we contacted corresponding authors of 35 handwashing impact evaluations published since 2012, targeting those included in recent systematic reviews,^1,18,19^ asking for cost data regarding the interventions they evaluated. For the cost of promotion, we identified 14 interventions (Supplementary Material B).^20,21,30,31,22–29^ For the cost of HWFs and annual expenditure on soap for handwashing, we identified 16 and 10 data points respectively. In addition, we were able to estimate the cost of formative research for five of the 14 promotion interventions, for which the 2019 US$ mean was about $100,000.^20–24^

For the cost of water for handwashing, we estimated an average annual cost separately for urban/rural areas. This was based on the proportion of the population using piped improved water supplies per country,^4^ and assumptions about the economic cost of water for piped and non-piped users based on average national tariffs, and water volumes used for handwashing (Supplementary Material B).^32^ This aims to represent the recurrent cost of water from existing supplies. The additional capital costs of providing new supplies to those who still lack improved drinking water sources were not included.

### Intervention definition

There are many possible approaches to promoting uptake of hand hygiene behaviours and facilities – no pre-existing intervention typology is universally used and no single programmatic approach is dominant.^19^ We aimed to estimate costs of an intervention representing “normative best practice”^12^ to the extent possible in this context. This was defined in consultation with an expert steering group and based on the types of interventions prevalent in the available price data. We characterised the intervention in the base case as a hygiene promotion campaign with modes of delivery including one-to-one promotion (e.g. house-to-house visits), group activities (e.g. community meetings, roadshows, street theatre) and mass media (e.g. radio, television, social). We assume that it is preceded by formative research to identify target drivers of behaviours and the development of a comprehensive behaviour change strategy.^33^

### Relation of intervention characteristics to effectiveness and price

We know of only one review exploring whether more intensive interventions have greater effects on health or behaviour. Pickering et al. undertook a systematic review of the impact of handwashing interventions on diarrhoea, focusing on the characteristics of studies which did and did not report a statistically significant reduction in diarrhoea.^34^ Of the 10 identified handwashing studies which reported frequency of contact between promoter and participant, none of the 3 which contacted participants once per month reported a significant effect on diarrhoea. Amongst the remaining 7 which contacted participants twice a month or more, 5 reported a significant effect on diarrhoea (of which 3 contacted twice a week or more). Similar findings were reported for point-of-use water treatment.^34^ It is not possible to say whether the additional cost of more intensive interventions justifies increased likelihood of health effects, since there is only one primary cost-effectiveness study of a hygiene intervention in a LMIC domestic setting.^20,35^

In support of intervention definition for our study, IR and JEM extracted data on characteristics of the 14 promotion interventions for which we had cost data, including: the nature of formative research; theoretical basis;^19^ modes of delivery; scale; and frequency (e.g. number of visits/meetings). Categorising interventions by these characteristics, we assessed the relationship between each characteristic and price per household (to the extent possible, given missing data). Other than scale, the design-relevant characteristic which appeared highly correlated with price per household was modes of delivery employed, particularly whether one-to-one promotion activities (e.g. household visits by a promoter) were included. Given the apparent higher effectiveness of more intensive interventions,^34^ we include one-to-one promotion in the intervention modelled in our ‘base case’. However, we also model a lower-cost ‘alternative case’ which excludes one-to-one promotion. We present box plots of the distributions of input price variables (Figure 1).

**Figure 1:**
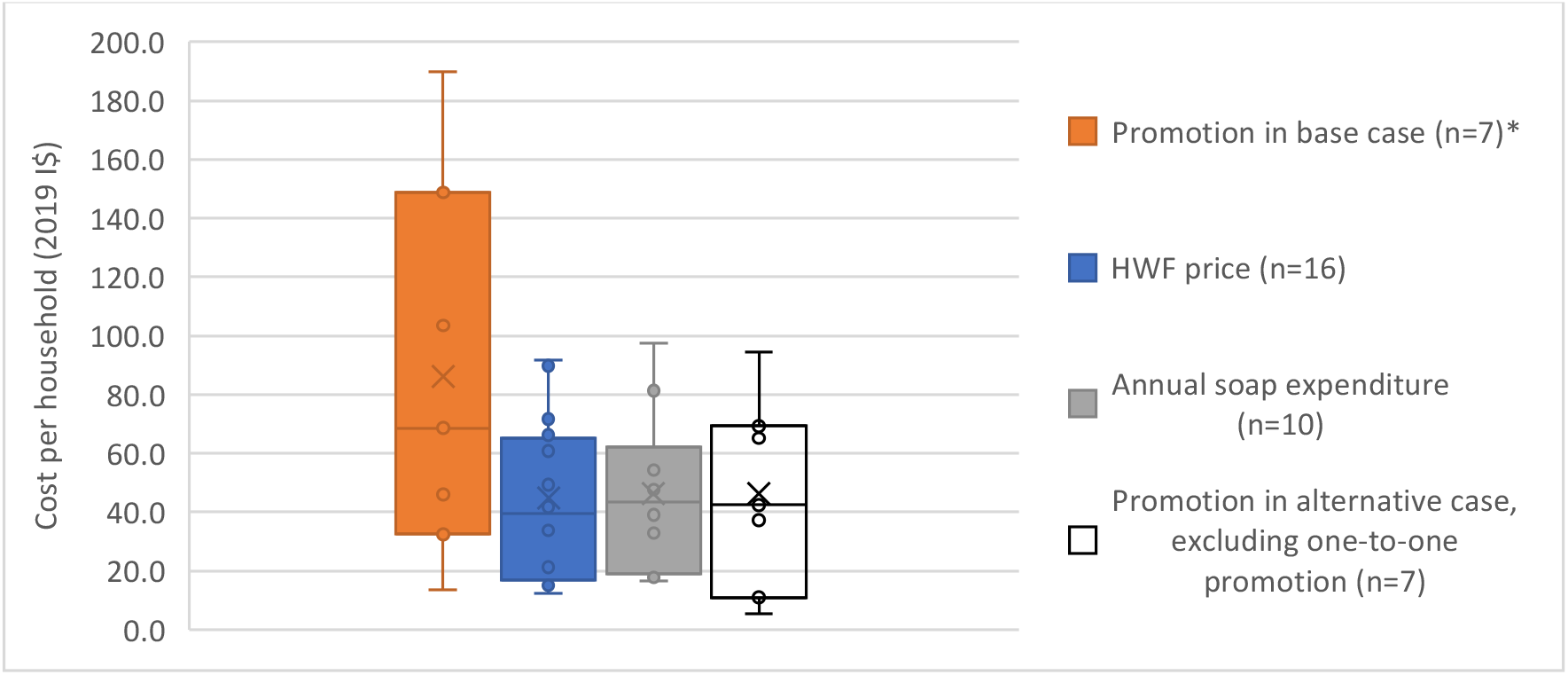
Distribution of key input price variables in 2019 international dollars (I$) * The base case promotion intervention includes one-to-one activities, group activities and mass media. The alternative case excludes one-to-one activities. This figure presents results in I$. Median US$ prices per household after converting to local purchasing power are $33.9 for base case promotion, $16.6 for HWF, $17.1 for soap expenditure, and $17.2 for alternative case promotion. Median annual expenditure per household on water for handwashing was $5.7 in rural areas and $4.0 in urban areas. The US$ median for the home-made HWF considered under sensitivity analysis was $1.2.

It was not possible to account for scale empirically, with too few datapoints for larger-scale interventions and the larger-scale interventions also tending to include fewer modes of delivery. In the base case, we applied the mean of prices from studies of interventions including one-to-one promotion, which represents interventions delivered at various scales. We model assumptions for economies of scale in sensitivity analysis.

### Cost categories

There are six cost categories (Table 1). Conceptualising promotion as a software capital investment, its useful life is assumed to be five years, after which it is repeated in full. “Top-up” promotion occurs annually, based on the theory that messages need to be repeatedly reinforced for behaviour change to be sustained. Following the intervention, all households are assumed to acquire a HWF. Thereafter, recurrent costs of supplying soap and water are incurred.

**Table 1:** Cost categories.

| Cost category | Description and key assumptions |
| --- | --- |
| **1. Formative research**<br>(software capital) | Design-focused research and piloting to identify target drivers of behaviours and the development of a comprehensive behaviour change strategy. |
| **2. Promotion**<br>(software capital) | Hygiene promotion campaign with a useful life of 5 years, and modes of delivery including: (i) house-to-house visits by promoters; (ii) community / group activities; and (iii) mass media (Supplementary Material B). |
| **3. Handwashing facility**<br>(hardware capital) | Purpose-built 20-litre drum with tap, basin and stand, with a useful life of 5 years. No capital maintenance is assumed, due to short useful life and very simple infrastructure. |
| **4. Top-up promotion**<br>(software capital maintenance) | Additional promotion activities occurring annually at 25% of initial cost (an assumption based on expert judgement), representing a lighter version of the intervention with lower frequency and dose |
| **5. Soap** (recurrent) | Expenditure on soap for handwashing (i.e. excluding other uses). The type of soap varied across studies but was predominantly bar soap. |
| **6. Water** (recurrent) | Expenditure on water used for handwashing, assumed to average 1.5 litres/person/day (Supplementary Material B).<sup>36</sup> |

### Statistical analysis and sensitivity analysis

We annuitised capital costs and discounted all costs at 3% in the base case following reference case guidance.^12,37^ We report results in 2019 US dollars (US$). We analysed prices by first converting to 2019 prices in local currency to adjust for inflation.^38^ We then converted to 2019 international dollars (I$), using World Bank data on GDP deflators and purchasing power conversion.^39^ After estimating the I$ mean and standard error (s.e.) per cost category for available data, we converted them to 2019 US$ per country. The rationale for converting to I$ to take the mean is that the US$ mean would be biased by purchasing power. For example, the I$ mean for annual soap expenditure in our data is $46, while the equivalent US$ value is $11 at Afghanistan’s purchasing power, or $20 in Niger.

All parameters in models are uncertain, but some are based on empirical data while others are the analyst’s assumptions. Interventions costed in the studies underlying our price estimates varied, e.g. in their modes of delivery, programme design, and scale. We sought to characterise parameter uncertainty using probabilistic sensitivity analysis, by taking a Bayesian approach using repeated draws from prior distributions constructed from available price data. We developed a probabilistic estimate of total cost using 1,000 draws in a Monte Carlo simulation, calculating a 95% uncertainty interval based on iterations’ percentiles. Price data for all cost categories were right-skewed so were modelled as gamma distributed, in line with usual practice.^40^ In the absence of published confidence intervals for JMP data, quantities were not varied probabilistically. We also undertook deterministic sensitivity analysis to explore the impact of uncertainty surrounding individual parameters and assumptions (scenarios in Supplementary Material C). We present tornado plots indicating the magnitude of changes in total cost when parameters are at high and low plausible values.

### Patient and public involvement

Public involvement was not undertaken as the study analysed secondary data from 46 countries.

## 3. Results

In the base case, the total economic cost of hand hygiene for all in household settings in 46 LDCs over 10 years was between US$ 12.2 – 15.3 billion (95% CI, point estimate 13.7 billion) (Figure 2). The cost of initial hand hygiene promotion was $334 million per year on average (24% of total costs), with a further $233 million per year for ‘top-up’ promotion (17%). The cost of handwashing facilities, a purpose-built drum with tap and stand/basin per household, was $174 million per year (13%). The cost of supplies was $497 million per year for soap (36%), and $127 million for water (9%).

**Figure 2:**
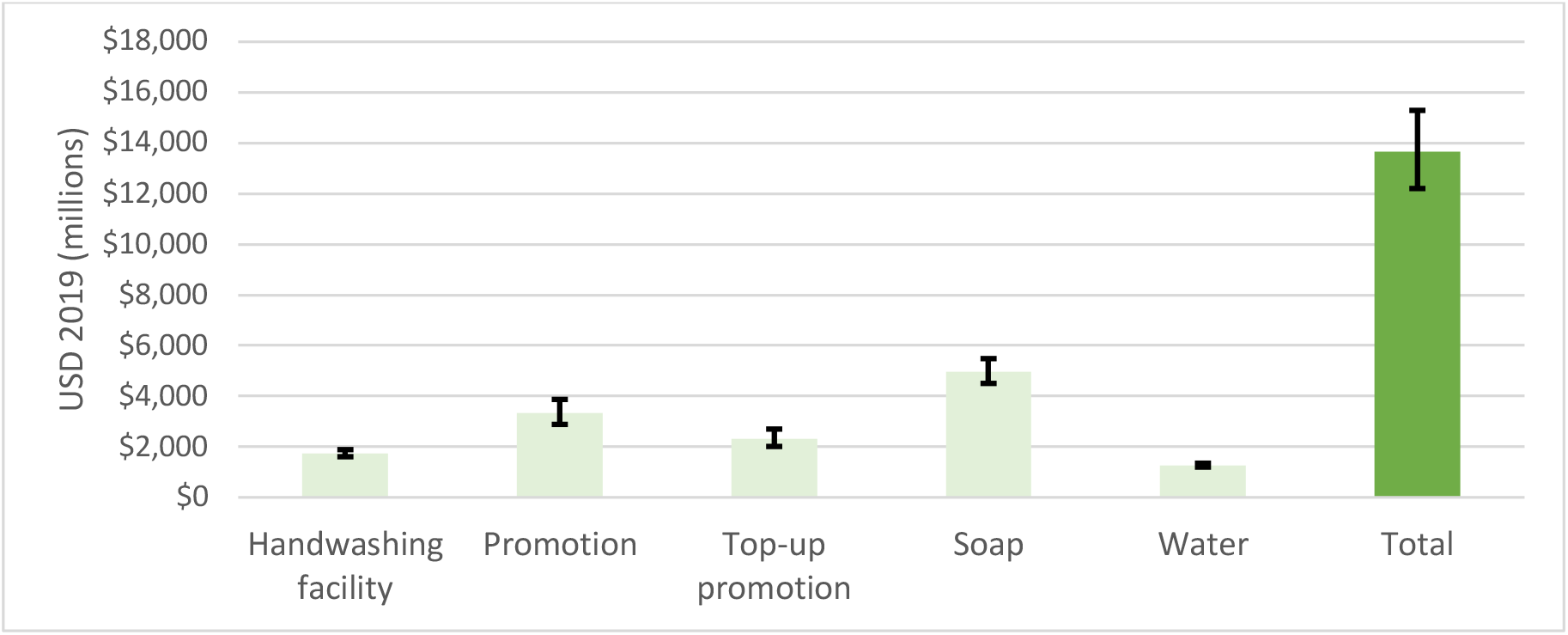
Total economic cost of hand hygiene for all in domestic settings in 46 least developed countries. *\* error bars represent the 95% confidence interval of 1,000 draws in Monte Carlo simulation. The cost of initial formative research ($5 million) is not shown*.

Considering only promotion costs, the total cost was $5.7 billion (95% CI: 4.9 - 6.6), or $0.47 annually per head of LDC population (42% of the total). Considering only HWFs and supplies, the total was $8.0 billion (95% CI: 7.3 - 8.7), or $0.66 annually per head of LDC population (58% of the total). An alternative framing of the facility and supply costs – considering cost per household – is an initial investment in a HWF (lasting 5 years) at a median of $17, accompanied by an annual cost of $17 for soap and $5 for water (Figure 1).

Different cost categories change in relative importance over time, as the intervention is scaled up over 10 years (Figure 3). Capital costs, specifically the initial promotion and HWF, are consistent over the years as each of the 10 cohorts receives the initial intervention – the slight annual decrease is due to discounting. Total recurrent costs increase every year, which is due to new cohorts receiving top-up promotion and spending on soap and water. Overall, capital costs comprise 37% of the 10-year total and recurrent costs 63%, recalling that top-up promotion is a recurrent cost (Table 1). In the alternative scenario where one-to-one promotion was excluded from the intervention, total cost decreased to 11.0 billion (95% CI: 10.0 - 12.2) and promotion cost to $3.0 billion (95% CI: 2.7 - 3.5), or $0.25 annually per head of LDC population – figures in Supplementary Material C.

**Figure 3:**
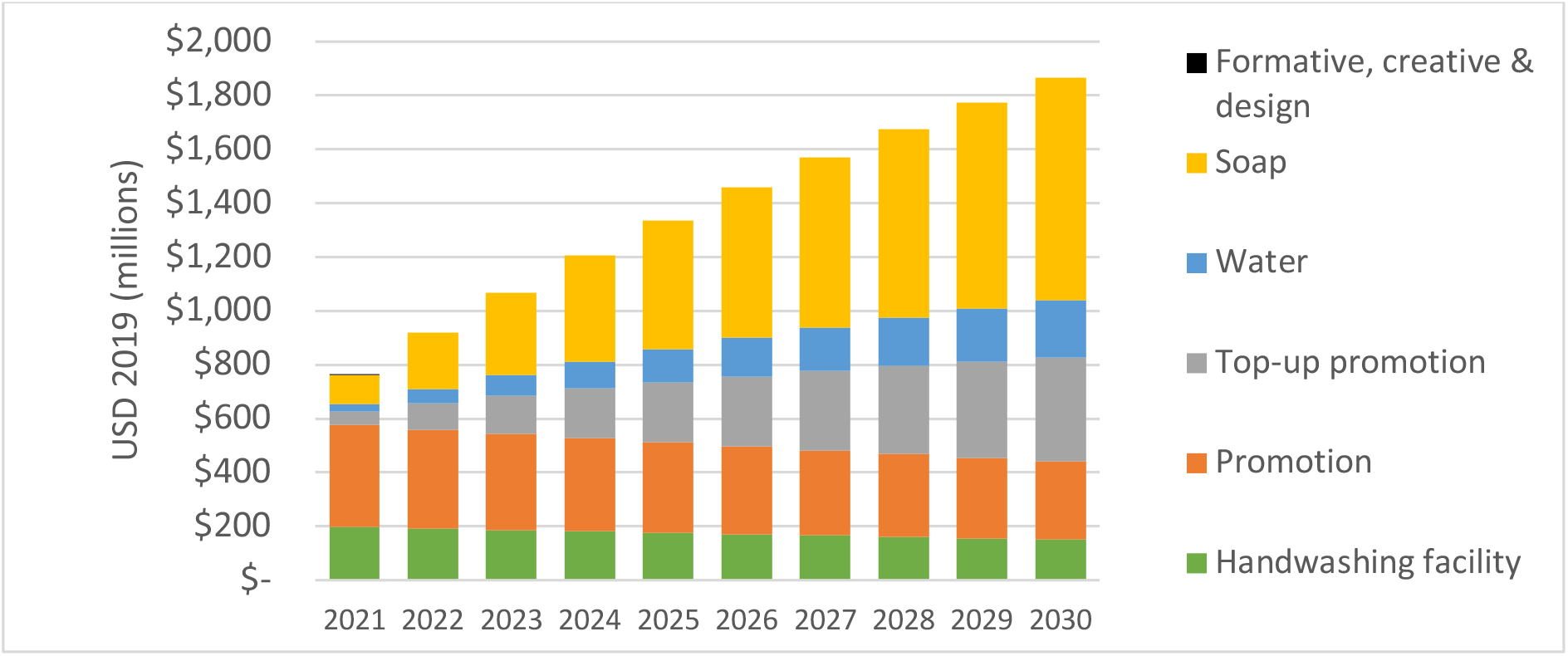
Distribution of costs over the 10-year time horizon, for 46 LDCs

Considering the first of the additional results, the annual cost of soap and water enabling households with a “limited” service to reach “basic” would be $1.4 billion, of which 86% is for soap. Second, if all households in LDCs were targeted with hygiene promotion instead of only those with no HWF, the promotion cost almost quadruples to $20.5 billion (from $5.7 billion) including one-to-one activities, and to $10.4 billion excluding them (from $3.0 billion). These amounts are not included in the headline totals.

In deterministic sensitivity analysis, the scenario which saw the biggest difference in total cost was when the promotion price was varied according to its 95% confidence interval. This saw total cost fall to US$ 10.5 billion or rise to $16.8 billion (Figure 4). Incorporating assumptions about economies of scale saw total cost falling to US$ 10.5 billion (Figure 4). In this scenario, the prices of promotion, HWFs and soap experience annual decreases starting with 10% in year 2 then in 2% decrements (8% in year 3, etc.). All scenarios are described in in Supplementary Material C, as are results for sensitivity analysis for promotion costs only.

**Figure 4:**
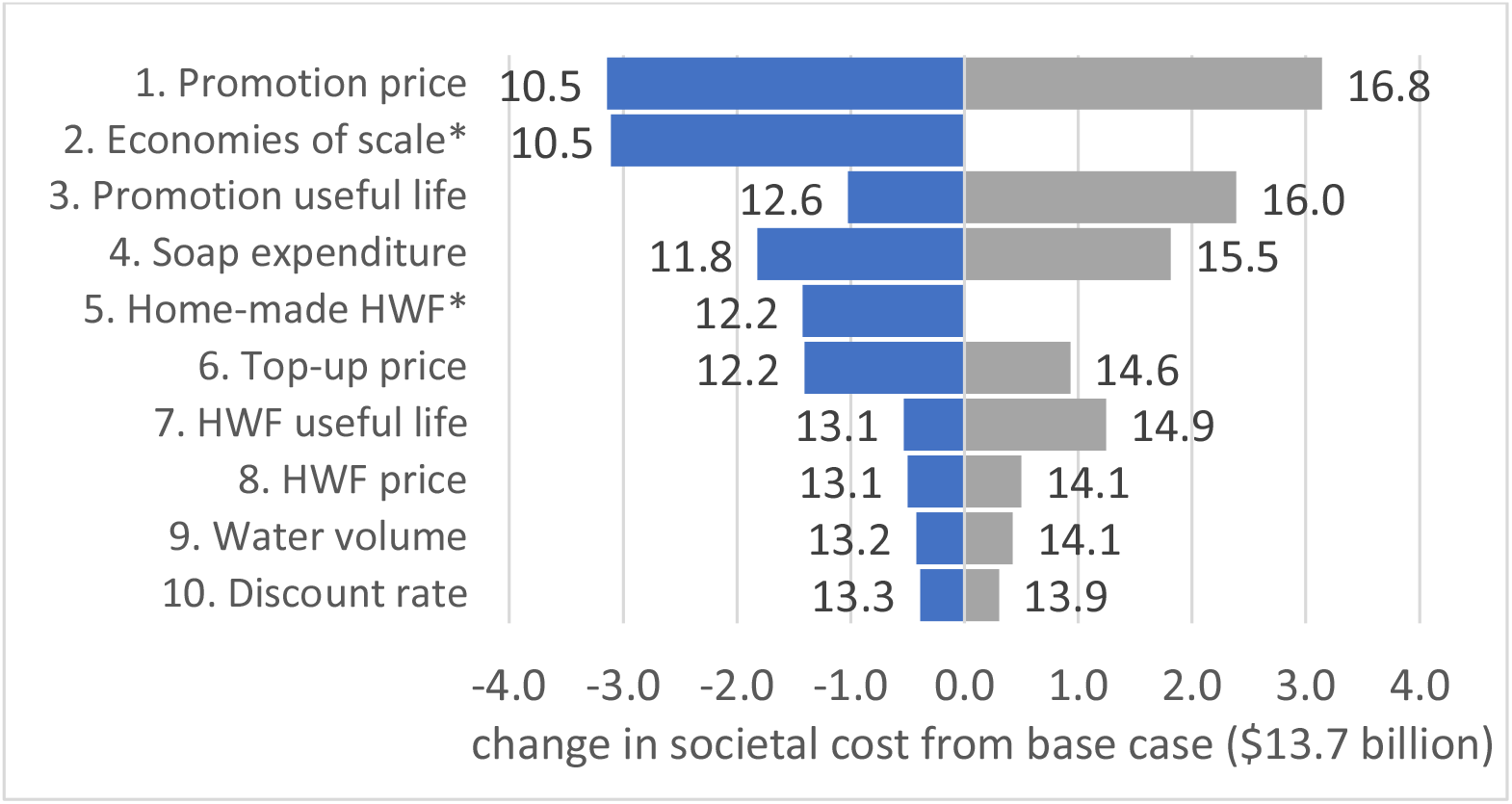
Tornado plot for total economic cost. note. bars represent lower and upper estimates of total societal cost when parameters are at high and low plausible values. Parameters with the largest bars contribute most to uncertainty. *no upper estimate

## 4. Discussion

Our study represents the most detailed to date of the costs of hand hygiene in domestic settings. We built on an earlier global study^10^ by differentiating between intervention modes of delivery and HWF technology through extracting study characteristics, by modelling alternative scenarios for these, and by characterising the uncertainty deriving from hygiene-related assumptions and price data. We also obtained far more price datapoints by contacting impact evaluation investigators and conducting new searches. Our application of probabilistic sensitivity analysis is a robust way of characterising uncertainty and heterogeneity in synthesising data from diverse interventions and contexts. Our study’s results can provide an empirical basis for country-level discussions of hand hygiene intervention prioritisation, costs, and how to cover them, which are increasingly taking place in light of the COVID-19 pandemic.^6^ To this end, WHO and UNICEF have developed an easy-to-use costing tool based on our model, which provides country-level estimates, and makes assumptions and input data easily editable by the user to reflect local realities.

Assuming that governments would fund promotion, top-up promotion, and formative research, the average annual cost was US$ 0.47 per head of LDC population. This represents 4.7% of median government health expenditure in LDCs ($10.0 per capita),^41^ and 12.3% of median WASH budgets of 23 LDCs ($3.8 per capita).^42^ The annual total represents 1% of the $57 billion in aid disbursed to LDCs by ‘official donors’ in 2019.^43^ Such expenditure may be justified on public health grounds since a substantial disease burden could be averted by hand hygiene,^3^ but better cost-effectiveness evidence would strengthen the case for health audiences. With only 16% of countries able to report the size of hygiene budgets or expenditures,^13^ wider application of the TrackFin methodology for estimating WASH expenditure would be beneficial.^44^ While the costs of HWFs, soap, and water are borne by households in the majority of cases, they can also be subsidised directly or indirectly (e.g. specific subsidies, cash transfers or humanitarian relief). The affordability of these costs for households requires investigation across a range of countries and settings.^45^ Soap presents a particular challenge, with survey data pointing to a strong socio-economic gradient in soap availability within households.^14^ The median annual cost per household of $17 for soap is likely to be unaffordable for the poorest households, and further innovation in handwashing technologies that reduce amounts of soap and water needed is required.^36,46^ Stimulation of soap markets in countries with particularly high prices would also be beneficial.

The headline hygiene-specific results of the previous global costing study^10^ are not directly comparable to our results, due to its reporting of hygiene-specific results for capital only and being for 140 countries. A comparison of unit prices is possible. Analysis of the earlier study’s raw price data^47^ indicates a median total capital cost of US$ 5-6 per person targeted (2015 prices), with software / promotion accounting for two thirds of the total.^48^ The median total capital cost in our study is US$9.9 per person targeted (2019 prices), with promotion accounting for 66% of the total. This suggests higher underlying prices, driven by: (i) our specification of a purpose-built HWF (when the earlier study’s price data included many basic types); and (ii) our inclusion of price data only for interventions including one-to-one promotion in the base case (when the earlier study pooled all types of intervention). The previous study’s median recurrent cost was $2-3 per person per year, which the underlying data suggest account for soap costs only. The equivalent for our study is $6.3 (of which soap accounts for 56%, water 19%, and top-up promotion 26%). Since water was accounted for separately in the previous study, not as part of the hygiene total, our higher recurrent cost estimate appears to be accounted for by having included top-up promotion.

Water is an important recurrent economic cost of handwashing regardless of whether it is paid for. Capital investment in water infrastructure will also be required to enable handwashing in many settings. In LDCs, 351 million people use a “less than basic” water supply, of which 141 million have a “limited” service comprising an improved infrastructure at greater than 30 minutes round trip.^4^ The less water there is easily available *in* the household, the lower the likelihood of handwashing taking place, which makes effective promotion harder.^7^ On-premises water supply that is available when needed is most likely to support handwashing, but only 37% of the LDC population have a “safely-managed” drinking water service that meets these criteria.^4^ Of the 81 million LDC households with “limited” hygiene, we estimated that 53% had no water available at the HWF at the time of survey. The World Bank estimated that universal “basic” water supply for those without it would cost US$ 7 billion per year and “safely managed” water supply $38 billion per year.^10^ Adding this to our results, to estimate the full cost of enabling hand hygiene, would substantially increase resource requirements.

We modelled scale-up as occurring in 10 equal cohorts over 10 years, but governments might decide to roll out interventions differently, for example as a national campaign like the Swachh Bharat Mission for sanitation in India.^49^ This would lead to more front-loading of costs, but likely also to economies of scale and scope. Our deterministic sensitivity scenario assuming annual reductions in unit prices for promotion, HWFs and soap (to a total of 30% on year 1 prices by the seventh year of implementation) sees total and provider costs reduce substantially (Figure 4 and Supplementary Material C). In addition, of the 14 promotion price datapoints, only 6 were for interventions reaching more than 10,000 households. Our input price data may therefore overestimate what it would cost to deliver interventions at large scale. Promotion price was the largest source of uncertainty (Figure 4), and the range of our input price distribution for promotion was very wide (Figure 1).

There may be efficiencies in combining handwashing messages with other messages, e.g. within WASH-specific programmes or broader programmes such as those delivered by health extension workers (HEWs).^50^ Of the time Ethiopian HEWs spend providing health education and services, they spend 30% of it on hygiene and environmental sanitation.^51^ However, this includes messaging on water management, faeces disposal, hand hygiene, food hygiene, waste management, etc., so actual HEW time spent on hand hygiene promotion is likely to be small. When messaging is diluted in this way there may be lower effectiveness than standalone campaigns, but this requires investigation. In addition, in some countries and settings community health workers are overstretched and lack the skills, training and support needed to deliver behaviour change programmes effectively.^52–54^

Following norms in resource requirement estimation,^12^ we estimate the cost of *reaching* all target households. Therefore, uptake and adherence are not accounted for, as would be necessary in a cost-benefit or cost-effectiveness analysis. As with many public health interventions, uptake of handwashing behaviours as a result of an intervention is typically below 50%, as is subsequent adherence.^19,55^ Even having a HWF with soap and water is no guarantee that hand hygiene is practised correctly at critical times, and promotion may benefit households already with a basic hygiene service.^5^ In the main analysis we costed interventions only for households with “no hygiene facility”. This was with a view to estimating the order of magnitude of costs, rather than suggesting that such households are specifically targeted by programmes. Since promotion costs almost quadruple if the whole population receives the intervention, there may be a balance to be sought. Some modes of delivery might be provided to whole populations, with others only being used in specific high-risk areas. For example, geo-referenced data from national health surveys could be used to identify sub-national areas with low HWF ownership or high cholera incidence.^56^

Taking into account the above considerations, the cost of achieving universal hand hygiene *behaviour*, as opposed to universal access to basic hygiene services, is likely to be larger than our headline estimate. Poor-quality cost evidence is a barrier to scaling up public health interventions and,^8,9^ with some notable exceptions,^20,30^ the costs of specific handwashing promotion interventions are rarely systematically collected. Impact evaluations need to collect intervention cost data if they are to effectively inform priority-setting,^57,58^ but we received usable cost data from fewer than half of the investigators contacted for this study. Almost all studies included in our model reported or necessitated top-down retrospective analyses, an approach with substantial limitations,^59^ and none included peer-reviewed studies employing bottom-up prospective costing.^60^ In addition, more studies and meta-analyses are required on the relative effectiveness of different approaches to hand hygiene promotion, which would also enable more intervention-specific costing.^19^

Limitations of our study are as follows. First, though we account for a higher economic cost of water for people without an on-premises piped supply, challenges of limited water availability for handwashing are not incorporated in any other way. Second, while we distinguish between hygiene service level differences between urban and rural areas, we apply a single national price per cost category, in the absence of reliable data on how prices of promotion, HWFs, soap etc. vary by urban/rural setting. Prices of many inputs are likely to be higher in rural areas further from markets, and income poverty is predominantly rural.^61^ Third, while we clearly specified the assumed intervention and extracted intervention characteristics from studies, the data underlying the sample mean for promotion price derive from many types of interventions at different scales.

## 5. Conclusion

We estimated the economic costs of universal access to basic hand hygiene services in household settings in 46 least developed countries, finding that US$ 12.2 – 15.3 billion is needed over 10 years. The promotion costs within this are $5.7 billion, representing 4.7% of median government health expenditure in LDCs, and 1% of their aid receipts. The remainder comprises HWFs ($1.7 billion), soap ($5.0 billion), and water ($1.3 billion). These costs could be covered by mobilising resources from across government and partners, and could be reduced by harnessing economies of scale and integrating hand hygiene with other behaviour change campaigns where appropriate. Innovation is required to make soap more affordable and available for the poorest households. Better evidence on the relative costs, effectiveness and cost-effectiveness of different promotion interventions for uptake and adherence of hand hygiene behaviours would enable more efficient investments.

## Data Availability

Quantity data are derived from public domain resources (http://washdata.org/). Price data are derived from published studies referenced in the manuscript. The economic model combining quantities and prices into cost estimates is available on request from the authors.

## Acknowledgements

We acknowledge the time and effort of impact evaluation investigators in answering our questions and providing data. We also acknowledge the advice and support of the steering committee, and seminar participants at LSHTM’s economic evaluation group and environmental health group.

## Author contributions

IR developed the methods with support from JEM/MM/RD. RJ/TS/GH provided data and analysis supporting methodological decisions and interpretation. IR searched for and extracted cost data. IR/JEM extracted intervention characteristics data. IR developed the model and analysed all data. All authors inputted into the manuscript.

