## Supplementary Materials for "Costs of hand hygiene for all in household settings - estimating the price tag for the 46 least developed countries"

### Online supplementary material

**A. Additional background**

- List of Least Developed Countries

**B. Additional methodological information**

- Model structure
- JMP imputation
- Formula for number of rural households with no hygiene service
- Electronic searches
- Intervention studies from which promotion price is derived
- Methods for estimating the cost of water

**C. Additional results**

- Results for alternative intervention scenario excluding one-to-one promotion
- Scenarios for deterministic sensitivity analysis
- Deterministic sensitivity analysis results for promotion cost

#### A. Additional background

*List of Least Developed Countries*

| **Americas and Caribbean (1)** | | **Middle East and North Africa (3)** | |
|---|---|---|---|
| Haiti | | Djibouti | Yemen |
| | | Sudan | |
| **East Asia and the Pacific (7)** | | **South Asia (4)** | |
| Cambodia | Solomon Islands | Afghanistan | Bhutan |
| Kiribati | Timor-Leste | Bangladesh | Nepal |
| Lao | Tuvalu | | |
| Myanmar | | | |
| **Eastern and Southern Africa (15)** | | **West and Central Africa (16)** | |
| Angola | Mozambique | Benin | Liberia |
| Burundi | Rwanda | Burkina Faso | Mali |
| Comoros | Somalia | Central African Republic | Mauritania |
| Eritrea | South Sudan | Chad | Niger |
| Ethiopia | Tanzania | Congo, D.R. | São Tomé and Principe |
| Lesotho | Uganda | The Gambia | Senegal |
| Madagascar | Zambia | Guinea | Sierra Leone |
| Malawi | | Guinea-Bissau | Togo |

#### B. Additional methodological information

##### *Model structure*

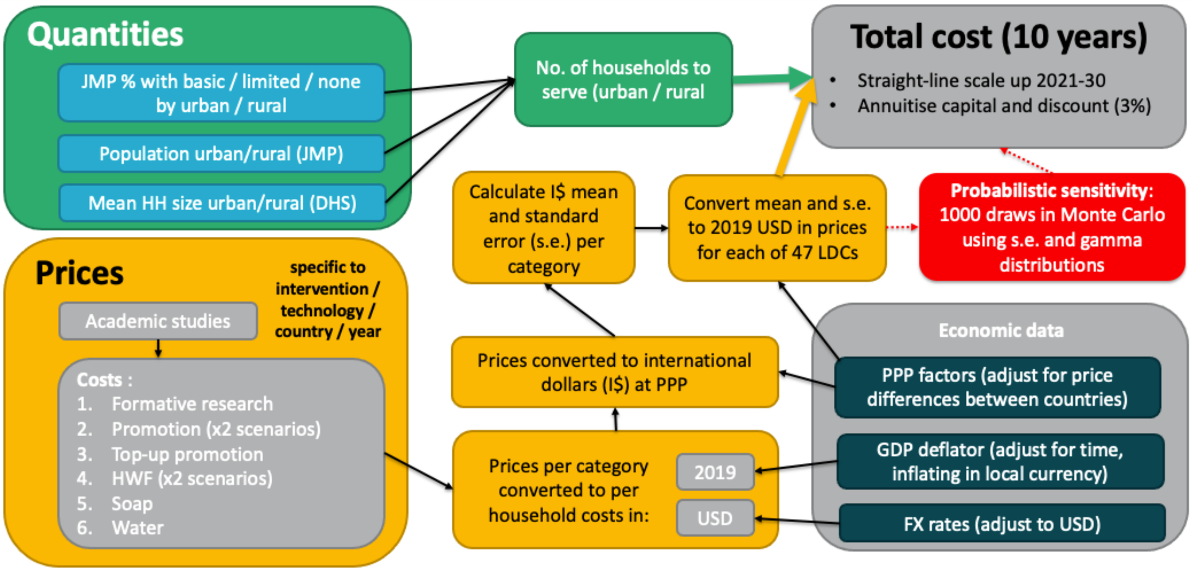


##### *JMP imputation*

The population of the five countries for which the LDC average is imputed represent only 6% of the total LDC population.

**Prior coverage year applied instead of 2020, in cases of missing data**

| Country | No hygiene facility | Piped / non-piped water supply |
|---|---|---|
| Comoros | 2016 | 2019 |
| Djibouti | 2020 LDC average* (21% urban, 30% rural) | 2020 |
| Eritrea | 2020 LDC average* (21% urban, 30% rural) | 2016 |
| Liberia | 2017 | 2020 |
| Mauritania | 2019 | 2020 |
| Mozambique | 2015 | 2020 |
| Solomon Islands | 2019 | 2020 |
| South Sudan | 2020 LDC average* (21% urban, 30% rural) | 2020 |
| Sudan | 2020 LDC average* (21% urban, 30% rural) | 2020 |
| Tuvalu | 2020 LDC average* (21% urban, 30% rural) | 2018 |
| Yemen | 2017 | 2020 |

*LDC average here means the average proportion of people across all LDCs with no hygiene facility

##### *Formula for number of rural households with no hygiene service*

We used the below formula to calculate numbers of households to be served in rural areas, per country. An equivalent formula was used for urban areas.

$$A_r = \frac{B_r}{C_r} * D_r$$

where:

$A_r$ is the number of rural households with "no hygiene service" in the country
$B_r$ is the total rural population in the country (UN-DESA medium variant 2019)
$C_r$ is the average rural household size in the country (latest DHS)
$D_r$ is proportion of the rural population in the country with "no hygiene service" (JMP data)

#### *Electronic searches*

On 4th June 2021, we searched Google Scholar for records since 2015, just before the Hutton & Varughese (2016) study was finalised. Search terms were handwashing cost (without inverted commas), "soap expenditure" and "expenditure on soap". We reviewed the first 10 pages of results for each search, downloaded full texts, and word-searched them for "cost", "$", "US", "price", and names/symbols of the currency of the study country.

#### *Intervention studies from which promotion price is derived*

Studies from which we extracted the price of hand hygiene promotion are listed below. Where source data excluded the costs of administration/management of the campaign, we attributed an uplift based on the average percentages for this cost from across studies that did so (24%).

**Promotion interventions (12 studies reporting 14 interventions)**

Borghi J, Guinness L, Ouedraogo J, Curtis V. Is hygiene promotion cost-effective? A case study in Burkina Faso. Trop Med Int Heal 2002; 7: 960–9.

Bikash Srot Kendra. Piloting hygiene promotion through routine immunisation in Nepal. 2017.

Delea MG, Snyder JS, Belew M, et al. Design of a parallel cluster-randomized trial assessing the impact of a demand-side sanitation and hygiene intervention on sustained behavior change and mental well-being in rural and peri-urban Amhara, Ethiopia: Andilaye study protocol. BMC Public Health 2019; 19: 1–15.

Briceño B, Chase C. Cost and Cost-Efficiency of Rural Sanitation and Handwashing Promotion: Activity-Based Costing and Experimental Evidence from Indonesia, India, Tanzania and Peru. 2014.

Pinfold J, Horan N. Measuring the effect of a hygiene behaviour intervention by indicators of behaviour and diarrhoeal disease. Trans R Soc Trop Med Hyg 1996; 90: 366–71.

Rajaraman D, Varadharajan KS, Greenland K, et al. Implementing effective hygiene promotion: Lessons from the process evaluation of an intervention to promote handwashing with soap in rural India. BMC Public Health 2014; 14. DOI:10.1186/1471-2458-14-1179.
Saadé C, Bateman M, Bendahmane DB. The Story of a Successful Public-Private Partnership in Central America. Handwashing for Diarrheal Disease Prevention. Arlington, Virginia: Basic Support for Child Survival Project (BASICS II), 2001.
Greenland K, Chipungu J, Curtis V, et al. Multiple behaviour change intervention for diarrhoea control in Lusaka, Zambia: a cluster randomised trial. Lancet Glob Heal 2016; 4: e966–77.
Evans B, Bates L, Halder A. Analysing the Value for Money of SHEWA-B in Bangladesh. 2015.
Waterkeyn J, Matimati R, Muringaniza A, et al. Comparative Assessment of Hygiene Behaviour Change and Cost-Effectiveness of Community Health Clubs in Rwanda and Zimbabwe. Healthc Access - Reg Overviews 2019. DOI:10.5772/intechopen.89995.
Biran A, White S, Awe B, et al. A cluster-randomised trial to evaluate an intervention to promote handwashing in rural Nigeria. Int J Environ Health Res 2020; 00: 1–16.
George CM, Monira S, Sack DA, et al. Randomized controlled trial of hospital-based hygiene and water treatment intervention (CHoBI7) to reduce cholera. Emerg Infect Dis 2016; 22: 233–41.

##### *Methods for estimating the cost of water*

Separately for urban and rural, we estimated a per country average price of water based on: (i) the proportion of households using piped improved water supply (JMP data for 2020);[1] (ii) the average national tariff per $m^3$ reported by the International Benchmarking Network for Water and Sanitation Utilities.[2] Households using piped improved were assumed to pay the IBNET tariff. Unconnected households were assumed to pay double that tariff, an approximation in the absence of data, to reflect their likely increased economic cost of water due to travel time. We combined the above prices with an assumed volume per person per day of 1.5 litres, to estimate an annual cost of water for handwashing. The volume estimate is based on: (i) an average of measured volume data reported by Whinnery et al.[3] for three types of barrel and tap technologies, tippy tap, and jug/basin; (ii) the assumptions that, in real life, people wash their hands for 10 seconds an average of four times per day.

#### C. Additional results

*Results for alternative intervention scenario excluding one-to-one promotion*

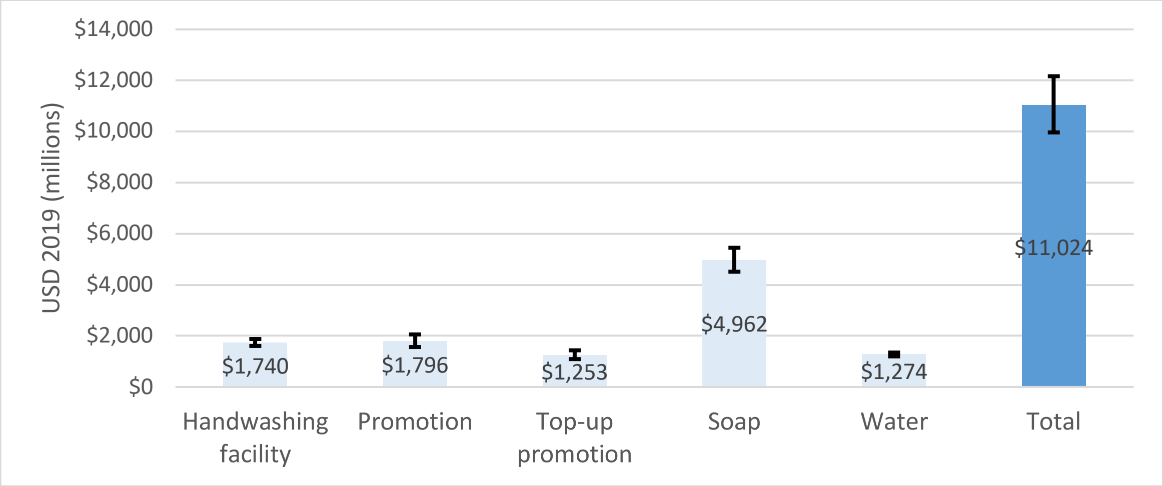


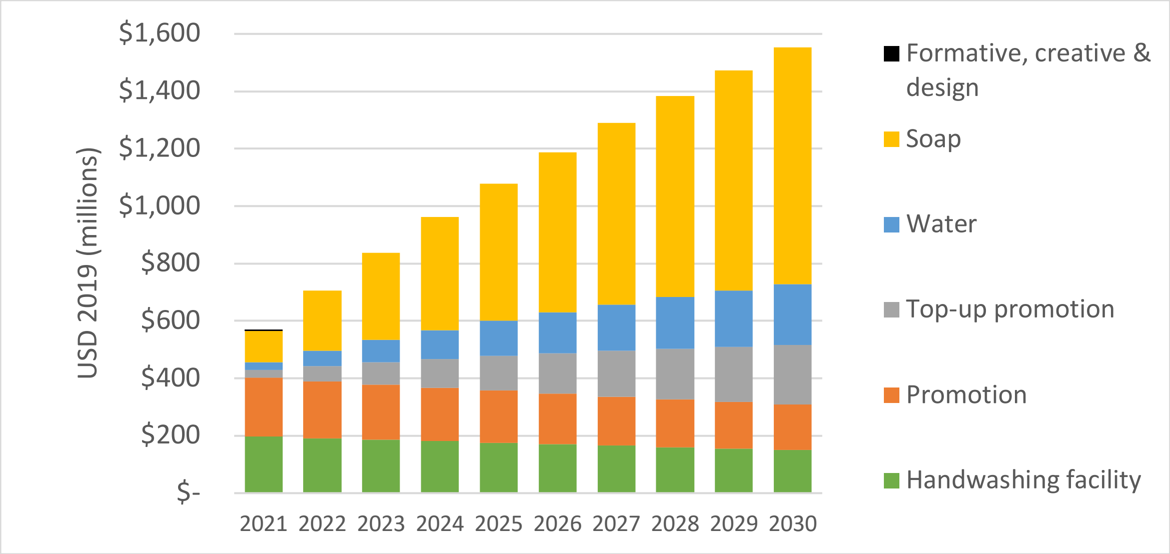

##### *Scenarios for deterministic sensitivity analysis*

| | | Variables | | |
|---|---|---|---|---|
| | | **Lower cost** | **Base case** | **Higher cost** |
| **Promotion** | Promotion price | lower bound of 95% CI (I$ 38) | mean (I$ 86) | upper bound of 95% CI (I$ 134) |
| | Top-up promotion price | 20% every 2 years | 25% every 1 year | 35% every 1 year |
| | Useful life of promotion | 7 years | 5 years | 3 years |
| **HWF** | HWF price | lower bound of 95% CI (I$ 32) | mean (I$ 45) | upper bound of 95% CI (I$ 58) |
| | HWF useful life | 7 years | 5 years | 3 years |
| | Home-made HWF instead of purpose built | “tippy-tap” or repurposed jug/bowl, with a useful life of 2 years (mean prices of n=4 studies) | purpose-built | n/a |
| **Other** | Annual soap expenditure | lower bound of 95% CI (I$ 29) | mean (I$ 46) | upper bound of 95% CI (I$ 63) |
| | Water volume used for handwashing (litres / person / day) | 1 | 1.5 | 2 |
| | Discount rate (based on IDSI/GHCC reference cases – see main body) | 7% | 3% | 0.1% |
| | Economies of scale | Prices of promotion, HWFs and soap in year 2 are 10% lower than year 1, further 8% lower in year 3, etc. such that price from year 7 onwards is 30% lower than year 1 & remains constant. | no change | n/a |

##### *Deterministic sensitivity analysis results for promotion cost*

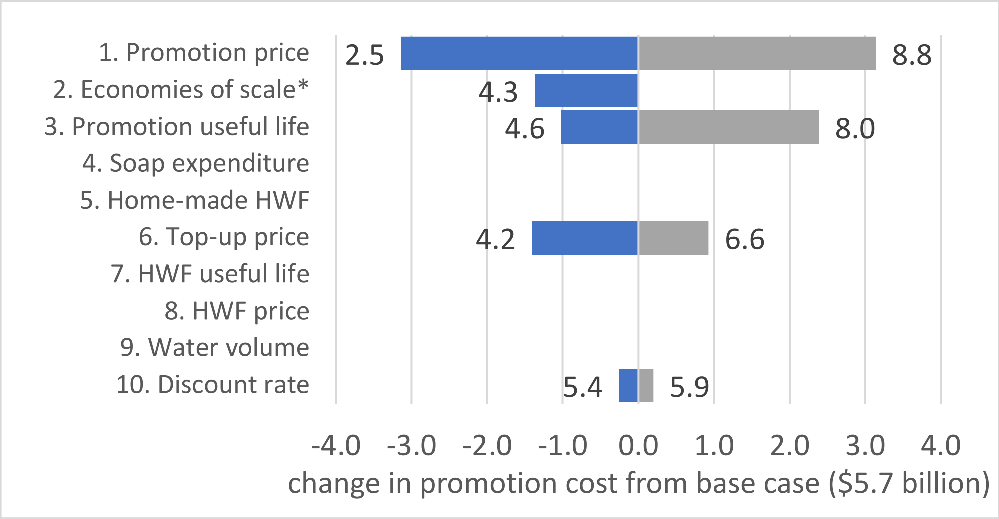
